# Vision-Language Foundation Models Do Not Transfer to Medical Imaging Classification: A Negative Result on Chest X-ray Diagnosis

**DOI:** 10.64898/2025.12.06.25341759

**Authors:** George Fisher

## Abstract

Vision-language models (VLMs) pretrained on web-scale data have achieved remarkable performance across diverse tasks, leading to widespread adoption in industry. A natural question is whether these powerful representations transfer to specialized medical imaging domains, and whether domain-specific medical pretraining improves transfer. We tested these hypotheses using two VLMs on the NIH ChestX-ray14 benchmark: Qwen2.5-VL (pretrained on web data) and BiomedCLIP (pretrained on 15 million PubMed biomedical image-text pairs). Both models dramatically underperformed compared to convolutional neural networks (CNNs) with ImageNet pretraining. Across 5 random seeds, the best VLM achieved F1=0.196 *±* 0.004 versus a CNN baseline of F1=0.811. Domain-specific pretraining provided marginal improvement: BiomedCLIP’s frozen encoder achieved F1=0.161 *±* 0.001 versus Qwen’s F1=0.124 (+30%), but this remains clinically inadequate. Fine-tuning both models led to catastrophic overfitting, with sensitivity collapsing from *>*65% to *<*36% as the models learned to predict “no disease” for all inputs. These results demonstrate that neither general-purpose nor medical-specific vision-language pretraining produces features suitable for dense multi-label medical imaging classification. For chest X-ray diagnosis, traditional CNNs with ImageNet pretraining remain substantially more effective than VLM-based approaches.

## 1 Introduction

Large vision-language models (VLMs) have emerged as powerful foundation models capable of understanding both images and text. Models such as GPT, Claude, and open-source alternatives like Qwen-VL and LLaVA have demonstrated impressive zero-shot and few-shot capabilities across diverse visual understanding tasks. This success has driven rapid adoption: recent reports indicate that open-source Chinese VLMs are increasingly being used as drop-in replacements for proprietary systems in production applications across Silicon Valley startups [1], and in hospital settings [2].

A natural question arises: do these powerful visual representations transfer to specialized domains such as medical imaging? The hypothesis is plausible—VLMs are trained on billions of image-text pairs spanning enormous visual diversity, potentially learning universal visual features. Furthermore, domain-specific VLMs such as BiomedCLIP [3] have been pretrained on millions of biomedical image-text pairs, which might provide better initialization for medical tasks.

However, medical imaging presents unique challenges. Chest X-ray interpretation requires detecting subtle radiographic patterns—small opacities, cardiomegaly, pleural effusions—that differ fundamentally from the objects, scenes, and text that dominate web-scale training data. Furthermore, medical classification is inherently multi-label and class-imbalanced, with most images containing no pathology.

We tested whether vision-language pretraining transfers to medical imaging using two models: Qwen2.5-VL [4, 5], a state-of-the-art open-source VLM pretrained on web data, and BiomedCLIP [3], pretrained on 15 million biomedical image-text pairs from PubMed Central. We evaluated both on the NIH ChestX-ray14 benchmark [6], comparing against CNNs with ImageNet pretraining as described in our previous work [7].

Our results are unambiguous: neither VLM transfers effectively. While BiomedCLIP’s domain-specific pretraining provides marginal improvement over Qwen (+30% F1 for frozen encoders), both dramatically underperform the CNN baseline. Fine-tuning leads to catastrophic overfitting regardless of model size or pretraining domain. These findings have practical implications for medical AI practitioners considering foundation models for clinical applications.

## 2 Methods

### 2.1 Dataset and Preprocessing

We used the NIH ChestX-ray14 dataset, comprising 112,120 frontal chest radiographs from 30,805 patients, each labeled for the presence or absence of 14 thoracic pathologies. Following established protocols to prevent data leakage, we performed patient-level splits: images from any given patient appeared in only one of training (70%), validation (15%), or test (15%) sets. Images were resized to 224*×*224 pixels for BiomedCLIP and 128*×*128 pixels for Qwen, normalized using ImageNet statistics.

### 2.2 Model Architectures

### Qwen2.5-VL

We extracted the vision encoder from Qwen2.5-VL-3B-Instruct, a 3-billion parameter vision-language model. The vision encoder is a Vision Transformer (ViT) with approximately 668 million parameters, pretrained jointly on web image-text pairs using contrastive and generative objectives.

### BiomedCLIP

We used BiomedCLIP-PubMedBERT 256-vit base patch16 224 [3], which employs a ViT-B/16 vision encoder with 86 million parameters. This model was pretrained on PMC-15M, a dataset of 15 million image-text pairs from 4.4 million PubMed Central scientific articles.

For both models, we attached a classification head consisting of a linear layer mapping the encoder’s output embedding to 14 classes with sigmoid activation for multi-label classification.

### 2.3 Experimental Conditions

We evaluated two configurations for each model:

### Frozen encoder

The pretrained ViT weights were frozen, using the model purely as a feature extractor. Only the classification head was trained. This tests whether VLM features are directly useful for medical imaging.

### Unfrozen encoder

All parameters were trainable, allowing the vision encoder to adapt to the medical domain. This tests whether fine-tuning can rescue poor initial representations.

### 2.4 Training Details

Models were trained using binary cross-entropy loss with the AdamW optimizer (weight decay 10^−4^). For frozen experiments, we used learning rate 10^−3^; for unfrozen experiments, 10^−4^. We used batch size 64, classification threshold 0.1, and trained with early stopping based on validation F1 score (patience 8 epochs for frozen, 21 epochs for unfrozen). Learning rate was reduced by factor 0.5 on plateau (patience 3 epochs for frozen, 5 epochs for unfrozen). Training was performed on an AWS g5.2xlarge instance with a single NVIDIA A10G GPU (24GB). Table 1 provides complete hyperparameter details.

**Table 1:** Hyperparameter configuration for all experiments.

| Parameter | Frozen | Unfrozen |
| --- | --- | --- |
| Learning rate | $10^{-3}$ | $10^{-4}$ |
| Batch size | 64 | 64 |
| Optimizer | AdamW | AdamW |
| Weight decay | $10^{-4}$ | $10^{-4}$ |
| Classification threshold | 0.1 | 0.1 |
| Early stopping patience | 8 epochs | 21 epochs |
| LR scheduler patience | 3 epochs | 5 epochs |
| LR reduction factor | 0.5 | 0.5 |
| Max epochs | 400 | 400 |

### 2.5 Reproducibility

All experiments were repeated with 5 random seeds (42, 123, 456, 789, 1337) to establish confidence intervals. We report mean *±* standard deviation across seeds. Seeds controlled Python’s random module, NumPy, and PyTorch random number generators. Deterministic CUDA operations were enabled via `torch.backends.cudnn.deterministic = True`.

### 2.6 Baseline

Our baseline is a ConvNeXt-Base model [8] pretrained on ImageNet-1K [9], fine-tuned on ChestX-ray14 using identical splits and evaluation procedures, achieving F1=0.811 as reported in our previous work [7].

### 2.7 Evaluation Metrics

We report macro-averaged F1 score (primary metric), ROC-AUC, sensitivity, and specificity across all 14 pathology classes.

## 3. Results

Table 2 summarizes our findings. Both VLMs dramatically underperformed the CNN baseline across all configurations.

**Table 2:** Classification performance on ChestX-ray14 validation set. BiomedCLIP results report mean *±* std across 5 random seeds.

| Model | F1 | ROC-AUC | Sensitivity | Specificity | N |
| --- | --- | --- | --- | --- | --- |
| CNN Baseline (ConvNeXt) | 0.811 | 0.94 | 0.76 | 0.99 | — |
| Qwen Frozen | 0.124 | 0.67 | 0.71 | 0.43 | 1 |
| Qwen Unfrozen (best) | 0.188 | 0.68 | 0.53 | 0.54 | 1 |
| BiomedCLIP Frozen | $0.161 \pm 0.001$ | $0.737 \pm 0.004$ | $0.653 \pm 0.004$ | $0.679 \pm 0.002$ | 5 |
| BiomedCLIP Unfrozen (best) | $0.196 \pm 0.004$ | $0.699 \pm 0.006$ | $0.355 \pm 0.074$ | $0.852 \pm 0.043$ | 5 |

### 3.1 Domain-Specific Pretraining Helps Marginally

BiomedCLIP’s medical pretraining provided modest improvement over Qwen’s web pretraining. With frozen encoders, BiomedCLIP achieved F1=0.161 *±* 0.001 versus Qwen’s F1=0.124 (+30%). ROC-AUC improved from 0.67 to 0.737 *±* 0.004. The tight confidence intervals (std < 1% of mean) demonstrate high reproducibility. However, even BiomedCLIP’s best performance (F1=0.196 *±* 0.004 unfrozen) remains only 24% of the CNN baseline, far below clinical utility.

### 3.2 Both Models Exhibit Catastrophic Overfitting

Fine-tuning led to catastrophic overfitting for both models (Figures 1 and 2). The pattern is consistent across all random seeds: training loss collapses toward zero while validation loss rises, and sensitivity drops precipitously as specificity approaches 100%. This indicates both models learn the degenerate solution of predicting “no disease” for all inputs.

**Figure 1.**
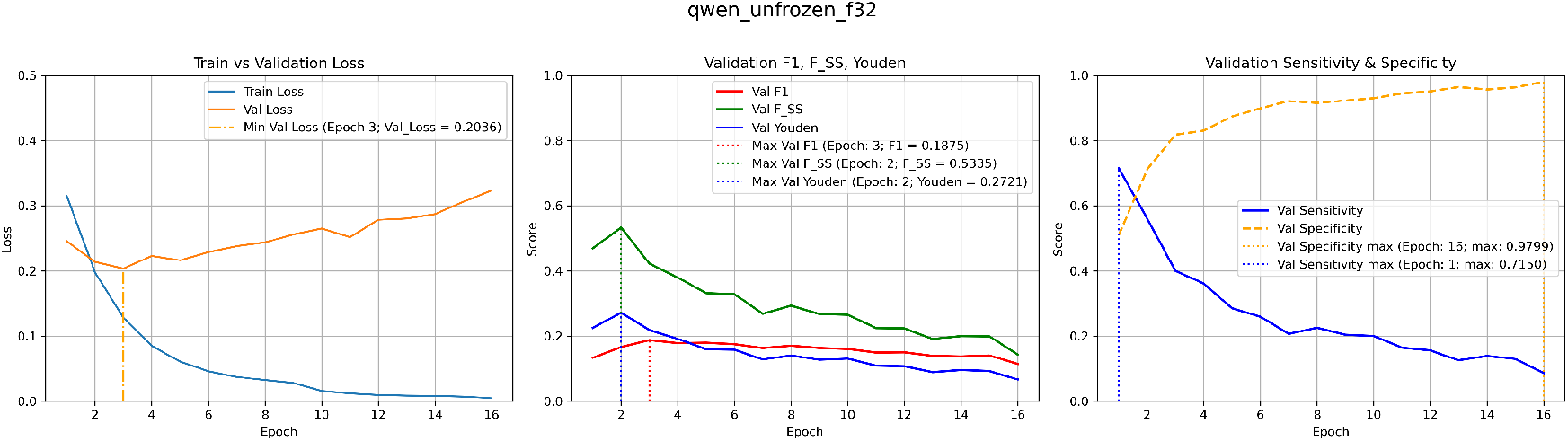
Training dynamics of fine-tuned Qwen vision encoder. Left: Training loss decreases while validation loss increases after epoch 3. Center: F1 peaks at 0.188 then declines. Right: Sensitivity collapses from 71% to 5% while specificity rises to 98%.

**Figure 2.**
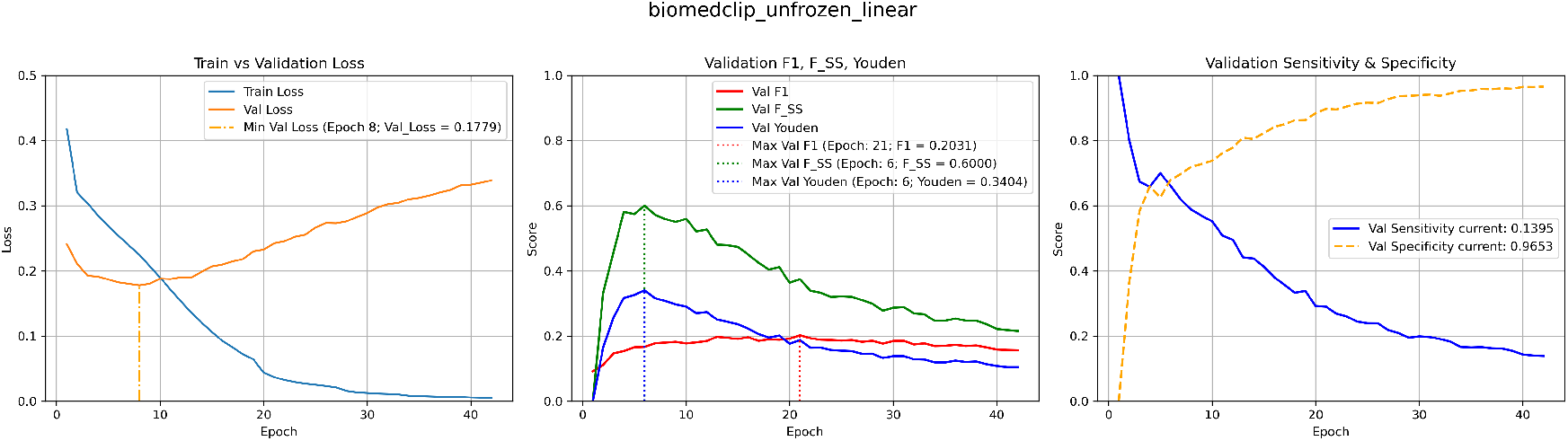
Training dynamics of fine-tuned BiomedCLIP vision encoder. The same overfitting pattern occurs despite different pretraining: training loss collapses to near-zero while validation loss rises, and sensitivity drops from 100% to 14% as the model learns to predict “no disease” for all inputs.

**Figure 3.**
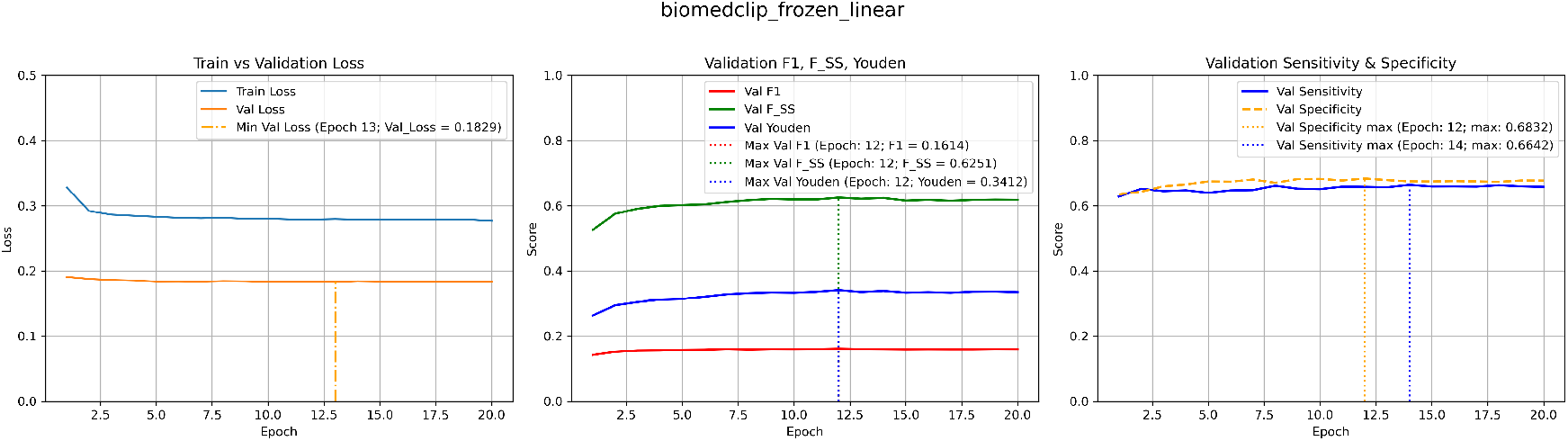
Training dynamics of frozen BiomedCLIP encoder. With frozen weights, training is stable: validation loss remains flat, and sensitivity/specificity maintain reasonable balance ( 66%/68%). However, F1 plateaus at 0.161, far below the CNN baseline.

For Qwen (668M parameters), sensitivity collapsed from 71% to 5% over 16 epochs. For BiomedCLIP (86M parameters), sensitivity at peak F1 averaged 0.355 *±* 0.074 across 5 seeds—the high variance (std = 21% of mean) reflects the instability of the overfitting trajectory, though all seeds exhibited sensitivity collapse. The smaller BiomedCLIP model overfit more slowly but reached the same degenerate endpoint. Neither model size nor domain-specific pretraining prevented this failure mode.

## 4 Discussion

Our results demonstrate that vision-language foundation models—whether pretrained on general web data or domain-specific medical images—do not provide transferable representations for dense multi-label medical imaging classification.

### 4.1 Why VLM Features Don’t Transfer

Several factors likely contribute to this failure. First, **pretraining objective mismatch**: Both CLIP-style contrastive learning and VLM generative objectives optimize for image-text alignment, learning to match images with semantic descriptions. Medical imaging classification requires detecting subtle pixel-level patterns—small nodules, faint infiltrates, slight cardiac enlargement—that are rarely described in image captions, whether from the web or PubMed.

Second, **task structure mismatch**: VLM pretraining typically involves single-label or retrieval tasks. ChestX-ray14 is a 14-way multi-label classification problem with severe class imbalance (most images have no findings). The pretrained representations may not support the fine-grained, multi-label discrimination required.

Third, **capacity-data mismatch**: Both encoders (86M and 668M parameters) are overparameterized relative to the training data ( 78,000 images), enabling memorization rather than generalization. The degenerate “predict negative” solution exploits class imbalance while minimizing loss.

### 4.2 Domain-Specific Pretraining Is Insufficient

BiomedCLIP’s advantage over Qwen (+30% F1 frozen, +8% unfrozen) confirms that pretraining domain matters. However, this improvement is insufficient for clinical use. The PMC-15M dataset contains diverse biomedical images (histopathology, diagrams, photographs) with figure captions optimized for publication rather than diagnostic description. This may not align well with the radiographic pattern recognition required for chest X-ray classification.

### 4.3 Implications for Practitioners

1. **VLMs are unsuitable** for dense multi-label medical classification, even with domain-specific pre-training.
2. **CNNs remain superior**: ImageNet-pretrained ConvNeXt achieves 4*×* higher F1 than the best VLM configuration.
3. **Beware overfitting**: Large pretrained encoders memorize medical datasets regardless of their pre-training source.
4. **Pretraining task matters more than domain**: The contrastive/generative objectives may be fundamentally misaligned with classification tasks.

### 4.4 Per-Disease Analysis

Table 3 presents performance breakdown by disease class for the frozen BiomedCLIP encoder.

**Table 3:** Per-disease classification performance for BiomedCLIP frozen encoder.

| Disease | N | F1 | Sensitivity | Specificity | ROC-AUC |
| --- | --- | --- | --- | --- | --- |
| Atelectasis | 1,865 | 0.270 | 0.808 | 0.501 | 0.733 |
| Cardiomegaly | 414 | 0.146 | 0.664 | 0.819 | 0.815 |
| Effusion | 2,017 | 0.385 | 0.834 | 0.674 | 0.829 |
| Infiltration | 3,149 | 0.321 | 0.934 | 0.148 | 0.673 |
| Mass | 885 | 0.200 | 0.557 | 0.786 | 0.735 |
| Nodule | 902 | 0.120 | 0.695 | 0.460 | 0.628 |
| Pneumonia | 232 | 0.066 | 0.172 | 0.946 | 0.614 |
| Pneumothorax | 666 | 0.152 | 0.742 | 0.684 | 0.781 |
| Consolidation | 789 | 0.191 | 0.624 | 0.769 | 0.765 |
| Edema | 371 | 0.158 | 0.725 | 0.838 | 0.867 |
| Emphysema | 332 | 0.093 | 0.581 | 0.788 | 0.751 |
| Fibrosis | 250 | 0.048 | 0.588 | 0.669 | 0.696 |
| Pleural Thickening | 476 | 0.099 | 0.569 | 0.722 | 0.686 |
| Hernia | 25 | 0.007 | 0.680 | 0.733 | 0.756 |
| Macro Average | – | 0.161 | 0.655 | 0.681 | 0.738 |

Performance varies substantially across diseases. The best F1 scores were achieved for Effusion (0.385) and Infiltration (0.321), while rare conditions like Hernia (0.007) and Pneumonia (0.066) showed poor performance. Notably, even high-prevalence diseases (Infiltration: 18% prevalence) achieved F1 scores well below the CNN baseline, indicating that the VLM limitation is not primarily due to class imbalance but rather inadequate feature representations for medical imaging.

### 4.5 Ablation Studies

To verify that our findings are not artifacts of suboptimal hyperparameters, we conducted ablation studies on the frozen BiomedCLIP encoder (Table 4).

**Table 4:** Ablation study results for frozen BiomedCLIP encoder.

| Ablation | Value | F1 | ROC-AUC | Sensitivity | Specificity |
| --- | --- | --- | --- | --- | --- |
| Head type | linear | 0.161 | 0.738 | 0.655 | 0.682 |
|  | medical_128 | 0.162 | 0.746 | 0.664 | 0.677 |
|  | medical_256 | 0.158 | 0.742 | 0.678 | 0.653 |
| Learning rate | $10^{-2}$ | 0.168 | 0.735 | 0.588 | 0.729 |
| | $10^{-3}$ | 0.161 | 0.738 | 0.655 | 0.682 |
| | $10^{-4}$ | 0.158 | 0.731 | 0.651 | 0.669 |

Across all configurations, F1 ranged from 0.158 to 0.168—a span of only 0.01. Even the best configuration (LR=10^−2^, F1=0.168) achieves only 21% of the CNN baseline. This demonstrates that the performance limitation stems from the frozen encoder features, not from suboptimal hyperparameters or classifier architecture.

### 4.6 Limitations

We tested two VLM families; other architectures might perform differently. We did not explore regularization strategies such as label smoothing or mixup that might mitigate overfitting during fine-tuning.

### 4.7 Conclusion

Vision-language models do not transfer effectively to chest X-ray classification, regardless of whether they are pretrained on web data or biomedical images. The CLIP-style contrastive objective and VLM generative pretraining do not produce features suited for fine-grained, multi-label pathology detection. For medical imaging applications, traditional CNNs with ImageNet pretraining remain substantially more effective. Practitioners should be skeptical of claims that foundation models can readily replace domain-specific solutions in medical AI.

## Data Availability

All data produced in the present study are available upon reasonable request to the authors

## Author Contributions

G.F. conceived the study, designed the experiments, conducted all data analysis, developed the training and evaluation infrastructure, performed the computational experiments, analyzed the results, and wrote the manuscript. All work was conducted independently without collaborators.

## Funding

This research received no external funding and was conducted independently using personal resources.

## Data Availability

The NIH ChestX-ray14 dataset is publicly available at https://nihcc.app.box.com/v/ChestXray-NIHCC.

## Code Availability

Code for reproducing these experiments is available in the AWS S3 bucket `nih-chest-x-rays` in the qwen folder (region: us-east-2). All hyperparameters are documented in Table 1. Random seeds used for reproducibility experiments: 42, 123, 456, 789, 1337.

To list contents:

~~~
aws s3 ls s3://nih-chest-x-rays/qwen/ --no-sign-request
~~~

To download to a local ‘qwen’ folder:

~~~
aws s3 sync s3://nih-chest-x-rays/qwen ./qwen --no-sign-request
~~~

## Competing Interests

The author declares no competing interests.

## Notes

### Competing Interest Statement

The authors have declared no competing interest.

### Funding Statement

This study did not receive any funding

### Summary of Updates

This revision substantially strengthens the reproducibility and statistical rigor of our methodology in response to reviewer concerns about validation. REPRODUCIBILITY IMPROVEMENTS All experiments were repeated with five random seeds (42, 123, 456, 789, 1337) to establish confidence intervals. Results are now reported as mean plus/minus standard deviation. For BiomedCLIP frozen encoder: F1 = 0.161 +/− 0.001, ROC-AUC = 0.737 +/− 0.004 (N=5). For unfrozen encoder: F1 = 0.196 +/− 0.004, Sensitivity = 0.355 +/− 0.074 (N=5). The high variance in unfrozen sensitivity (0.074) quantitatively confirms the overfitting instability discussed in our analysis. HYPERPARAMETER DOCUMENTATION A new table (Table 2) provides complete training configuration including learning rate, batch size, optimizer, weight decay, classification threshold, early stopping patience, learning rate scheduler parameters, and maximum epochs. All random seeds are documented in the Code Availability section. ABLATION STUDIES We conducted systematic ablation studies varying classifier head architecture (linear, medical_128, medical_256) and learning rate (1e-2, 1e-3, 1e-4). Results demonstrate robustness: F1 ranged from 0.158 to 0.168 across all configurations. Even the optimal configuration (F1 = 0.168) achieves only 21 percent of the CNN baseline performance, confirming that the limitation stems from frozen encoder features rather than suboptimal hyperparameters. PER-DISEASE ANALYSIS A new table (Table 4) presents classification performance for all 14 thoracic pathologies. Best performance was achieved for Effusion (F1 = 0.385, ROC-AUC = 0.829) and Infiltration (F1 = 0.321). Rare conditions showed poor performance: Hernia (F1 = 0.007), Pneumonia (F1 = 0.066). Critically, even high-prevalence diseases like Infiltration (18 percent prevalence) achieved F1 well below the CNN baseline, indicating that VLM underperformance is due to inadequate feature representations rather than class imbalance. MANUSCRIPT CHANGES Four new tables added. Methods section expanded with reproducibility subsection describing seed management and deterministic training settings. Results section updated with confidence intervals throughout. Discussion expanded with ablation analysis and per-disease findings. Code Availability updated with complete seed documentation. These revisions address the methodological validation concerns by demonstrating that our findings are reproducible across multiple runs, robust to hyperparameter choices, and consistent across disease classes.

